# IVERMECTIN REPROPOSING FOR COVID-19 TREATMENT OUTPATIENTS IN MILD STAGE IN PRIMARY HEALTH CARE CENTERS

**DOI:** 10.1101/2021.03.29.21254554

**Authors:** Rossana Elena Chahla, Luis Medina Ruiz, Teresa Mena, Yolanda Brepe, Paola Terranova, Eugenia Silvana Ortega, Guillermo Gabriel Barrenechea, Daniel Gustavo Goroso, Maria de los Angeles Peral de Bruno

## Abstract

**Background:** The emergence of COVID-19 requires alternative treatments based on the reuse of drugs as a strategy to prevent the progression of the disease in patients infected with SARS-COV-2. The goal was to evaluate the use of ivermectin in mild stage outpatients to heal and / or reverse the progression of COVID-19 disease towards the development of moderate or severe stages.

**Methods:** Cluster Assigned Clinical Trial (2:1) in outpatients, n = 234. The subjects were divided into experimental (EG: n = 110) and control groups (CG: n = 62). The EG received ivermectin orally 4 drops of 6 mg = 24 mg every 7 days for 4 weeks. All participants were diagnosed by positive RT-PCR for COVID-19 and were evaluated by clinical examination, at the beginning and the end of protocol. Data analyzed were applied the proportion, bivariate, and logical regression tests with level significance p < 0·05. This study was registered at ClinicalTrials.gov Identifier NCT04784481.

**Findings:** Both groups were similar in age, sex, and comorbidities (EG: 56F, median age= 40·0, range: 18·0 - 75·0; CG: 34F, median age = 37·5, range: 18·0 - 71·0). A significant reduction in the symptom numbers was observed in the EG when the medical examination was performed from 5^th^ to 9^th^ days, after starting treatment (*p* = 0·0026). Although, medical examination from 10^th^ to 14^th^ day, showed a progressive reduction of the percentage symptom numbers, these were not significative in both groups. A higher proportion of medical release was observed in EG (98·2%) vs CG (87·1%) (*p* = 0·003). EG showed 8 times more chance of receiving medical release than CG (OR 7·99, 95% CI: 1·64 −38·97, *p* = 0·003). The treatment effect with ivermectin to obtain medical release was analyzed by the logistic regression model based in the following control variables: sex, age, and comorbidities. Then, the chance to obtain medical release was maintained in EG (OR 10·37, 95% CI: 2·05 - 52·04, *p* = 0·005).

**Interpretation:** Treatment with ivermectin in outpatients with mild stage COVID-19 disease managed to slightly reduce the symptom numbers. Also, this treatment improved the clinical state to obtain medical release, even in the presence of comorbidities. The treatment with ivermectin could significantly prevent the evolution to serious stages since the EG did not present any patient with referral to critical hospitalization.

*Clinical Trials* registry number is NCT04784481

**Funding:** Ministry of Public Health. Tucumán, Argentina.

**Research in Context:** *Evidence before this study:* Currently there are no specific therapies approved for COVID-19 treatment by the FDA, that is why different repositionable drugs are being studied in clinical trials and compassionate use protocols based on *in vitro* activity. ivermectin is a broad spectrum antiparasitic agent that has been shown to have antiviral activity against a wide range of viruses. A study by Caly et al. (2020) suggested thatnuclear transport inhibitory activity of ivermectin may be effective against SARS-CoV-2. Since the publication of that work, numerous clinical trials were started to study ivermectin potential for COVID-19 treatment. At the end of March 2021, there were about 60 studies registered in https://www.clinicaltrials.gov, and 43 studies listened https://www.who.int/clinical-trials-registry-platform about the safety and effectiveness of ivermectin in COVID-19 patients, for treatment and prophylaxis. Most of these studies are from developing countries, which shows the need of emerging economies to find alternative therapies to contain the spread of the disease and the collapse of health systems.

*Added value of this study:* We found that an early intervention with ivermectin has impacted on the score of symptoms related to COVID-19 in ambulatory patients, between the 5^th^ and 9^th^ day. The patients who received the treatment changed from score 2 to score 1 in the WHO ordinal scale. In any case, patients evolved to higher scores. Also the treatment increased the probability to obtain medical release, even in the presence of comorbidities.

*Implications of all available evidence:* According to the <u>COVID-19 Treatment Guidelines by the NIH</u>, most trials have several limitations. It needs results from adequately powered and well-designed clinical trials to provide evidence-based guidance on the role of ivermectin in the treatment of COVID-19. However, our study shows overlaps in benefits with other authors, and taking together, these results are encouraging for further study about repurposing ivermectin for the treatment of COVID-19.

## 1. Introduction

Since the beginning of the pandemic, despite drastic containment measures, the spread of this virus has threatened to collapse health systems around the world, and also had devastating socio-economic consequences worldwide^1^. On the other hand, the impact of the coronavirus pandemic was unequal considering the vulnerability of developing countries whose economies are less able to cope with the new challenges imposed^2^.

International health authorities have focused on the rapid diagnosis and isolation of patients, as well as the search for therapies capable of counteracting the most serious effects of the disease, which constitute approximately 15% of the cases accounting to the World Health Organization (WHO)^3,4^. As the number of infected increases exponentially, the development of vaccines and new antiviral therapies becomes urgent. Unfortunately, these developments are outside the current timeline to contain the pandemic. In this context, the repositioning of drugs currently available on the market with established safety profiles that are implemented on another therapeutic indication, based on solid preclinical studies, is imperative. This is a pragmatic strategy, a faster and cheaper option compared to the new drug development, that has proven successful for many medicaments, and can be a key tool in emergency situations such as the current one that requires a quick action^5–8^. This strategy becomes more relevant in those economies that do not have the necessary resources for the development of new therapies, as in the case of Latin-American countries.

Ivermectin is a broad spectrum antiparasitic agent that has been shown to have antiviral activity against a wide range of viruses and there are studies which propose it as a candidate for COVID-19 treatment. Caly et al. (2020) suggested that ivermectin’s nuclear transport inhibitory activity may be effective against SARS-CoV-2^9^. In line with this study, numerous clinical trials, especially from developing countries, are evaluating the potential of ivermectin against COVID-19 with results that are not conclusive yet regarding its efficacy and safety^10–12^.

In our health system, the emergency of COVID-19 requires the urgent development of strategies to avoid the impact of the disease on our population, the saturation of the health system and that allows us to carry out adequate treatments to reduce the mortality of the disease. In this context, our health system considered the study of the repositioning of ivermectin as a strategy to stop and / or reverse the progression to developing moderate or severe stages of COVID 19 disease since: a) it is a safe drug, b) available in our environment c) with preclinical evidence of prophylactic capacity and d) antecedents in other health systems in the world of its use both preventively and in empirical treatment. We propose the use of ivermectin as one of the main pharmacological options whose repositioning was proposed for the therapeutic intervention of SARS-CoV-2.

The present study evaluates the use of ivermectin in mild-stage patients to heal and / or reverse the progression of COVID-19 disease towards the development of moderate or severe stages, using the ordinal scale of 8 points for therapeutic evaluation for COVID-19 recommended by the WHO^13^.

### *Primary Outcome* Decrease the number of patients with mild symptoms

*Secondary Outcomes*. Avoid progression of the disease to moderate or severe states until the end of the study, according to the 8-category ordinal scale used by WHO^13^.

## 2. Material and Methods

### 2.1. Sample Size

Sample size was determined by the test comparing two proportions^14^. It considered the following parameters to bilateral test: N = 30,000 Total operating area population for services network to Primary Care; 95% confidence level, 4% precision, 5% proportion, n = 148 sample size without loss, 15% expected proportion of losses. The sample size calculated was n = 174 participants.

### 2.2 Participants

The total group n = 240 enrolled outpatients from the urban area-department heads and peri-urban area of the City San Miguel de Tucumán. The study was conducted between September 2020 to January 2021. The health coverage service was administered by the Health System of the State of Tucumán (SI.PRO.SA, Tucumán, Argentina). The people who agreed to participate in the study gave their informed consent before starting the study (Research Ethics Committee / Health Research Directorate, file number 054/2020). The clinical trials registry number is NCT04784481. This study conforms to all CONSORT guidelines and reports the required information accordingly (see Supplementary Checklist).

### 2.3. Inclusion criteria

– Over 18 years of age of any sex.
– Outpatients infected by SARS-CoV-2 confirmed either by RT-PCR test.
– Women of childbearing age with a negative pregnancy test.
– Patients framed in the definition criteria of mild stage, that is; confirmed presence of two or more of the following symptoms: Fever less than 38·5°C, isolated diarrheal episodes, hyposmia or hypogeusia, mild desaturation (between 96 and 93%), dyspnea, polyarthralgia, persistent headache, abdominal pain, erythema of the kidney, nonspecific rash.

### 2.4. Exclusion Criteria

– Hypersensitivity or allergy to ivermectin.
– Pregnant or lactating.
– Children or adolescents under 18 years of age.
– Patients with neurological pathology, renal insufficiency, hepatic insufficiency.
– Weight less than 40kg.
– Patients with concomitant use of drugs that act on GABA, barbiturate and benzodiazepine receptors.
– Patients who have not completed / signed the informed consent.

### 2.5. Design

Clinical trial assigned by groups 2:1. The conglomerate of outpatients belonging to the urban area of county in the inside Tucumán was assigned to the Experimental Group (EG) and the outpatients from the peri-urban area of San Miguel de Tucumán and Gran San Miguel de Tucumán were assigned to the Control Group (CG). The criteria for this choice were based on the geographical distribution of Health Services, and logistics in current times of pandemic. Patients in urban areas in the interior of the province of Tucumán have poorer access to health than those who residing in the capital. This response to the unequal distribution of services and resources for most vulnerable sectors of society. The 2:1 ratio obeyed the criterion that the greatest number of outpatients received the intervention protocol. Figure 1 shows the Consort flow diagram.

**Figure 1.**
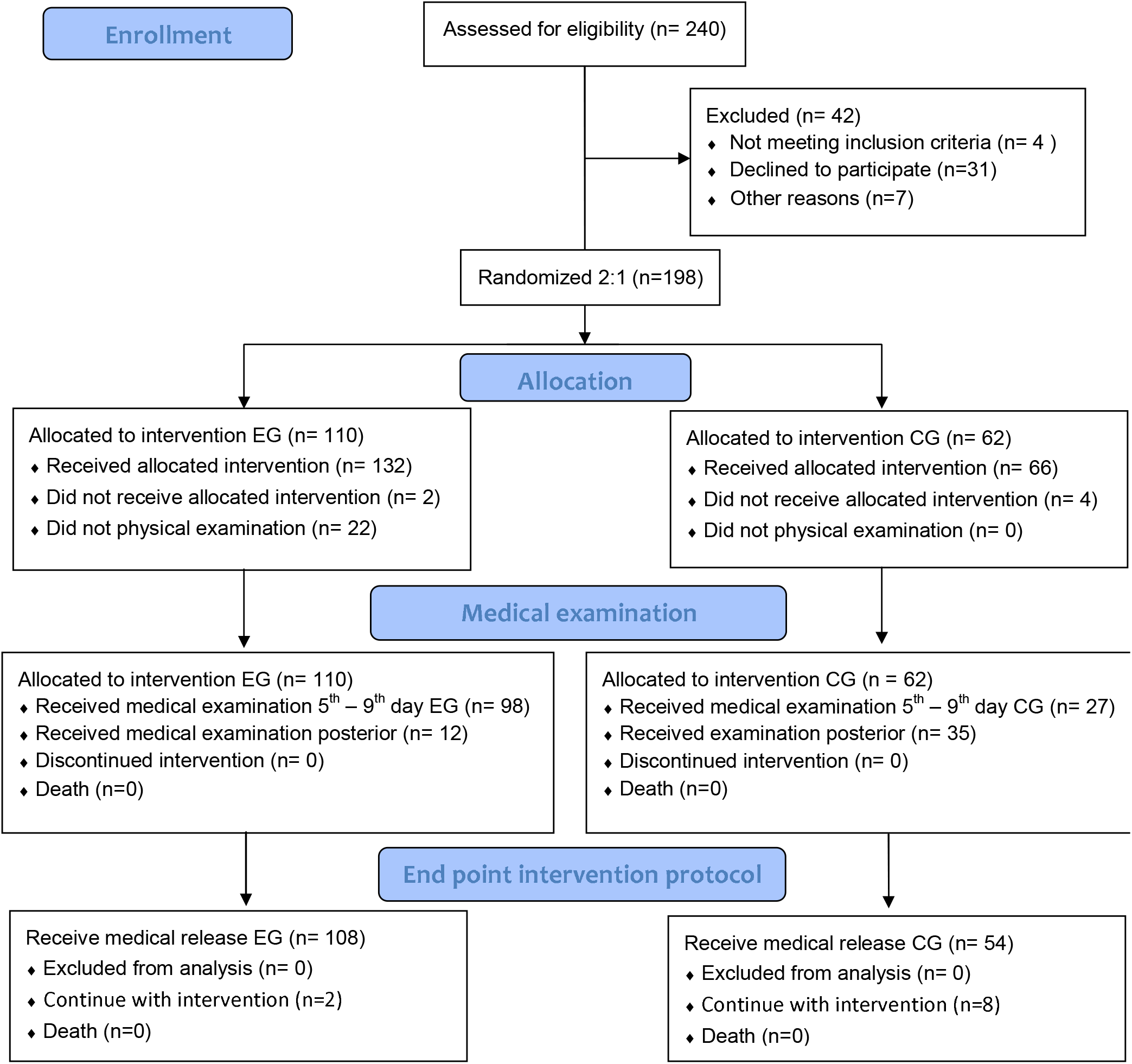
CONSORT flow diagram.

Figure 1 shows the flow that the patients followed during the treatment period. Note that 198 patients were initially recruited, allocated in a 2:1 ratio, GA (n = 110), and CG (n = 62). The 2:1 ratio obeyed the criterion that the greatest number of outpatients received the intervention protocol. The medical examination considered for a 1^st^ time frame was carried out from 5^th^ to 9^th^ day, this is since the effects of the treatment can manifest around 7 days (±2), being evaluated in this instance in EG (n = 98) and CG (n = 27). Other patients were reviewed in the 2^nd^ time frame: from 9^th^ to 14^th^ day EG (n = 12), and CG (n = 35). It should be noted that no deaths were reported and no patient left the intervention. Moreover, most of the participants were discharged from EG (n = 108), and for CG (n = 8), the rest continued under treatment and observation.

### 2.6. Intervention Protocol

The <u>EG received protocol ivermectin</u> orally 4 drops of 6 mg = 24 mg every 7 days for 4 weeks plus symptomatic treatment (500mg paracetamol every 6 or 8h, no more than 4 tablets daily; 100mg aspirin, 1 tablet per day with breakfast; 150mg Ranitidine, 1 tablet in the morning, and 1 tablet at night). By the other hand, CG received only symptomatic treatment. Patients with comorbidities continued with the basic medication for the underlying pathology. All participants were evaluated by physical examination COVID-19 diagnosed with positive RT-PCR at the beginning, and end of the protocol. A medical examination was carried out from 5^th^ and 9^th^ day (1^st^ time frame), and other outpatients were evaluated from 9^th^ to 14^th^ day (2^nd^ time frame). Enrolled subjects completed symptom questionnaires (including reporting of any adverse effects of treatment), physical examinations, and received medical release 4 weeks after the start of the intervention, and remote clinical telemedicine follow-up.

The 8-category ordinal scale recommended by the WHO was used to classified patients according clinical state, departing from category 2 of the scale at enrollment (not hospitalized and with limitation of activities)^13^. The patient status uninfected have a score 0, whom descriptor is no clinical or virological evidence of infection. This criterium was adopted to medical release.

### Security definitions

Adverse Event (AE) was defined as any medical event, signs, symptoms, or disease temporarily associated with the use of the medication, which could occur in the subjects enrolled in the study^15^.

### Adherence to treatment

The World Health Organization (WHO) defines adherence to treatment as compliance with it; that is, taking the medication according to the dosage of the prescribed schedule; and persistence, taking the medication over time^16^.

### Statistics

Categorical variables were analyzed with frequencies and percentages, and continuous variables with median and quartiles. Differences between the variables were determined using the Chi-square test. The proportion test and logistical regression were applied to calculate the subject’s proportions with symptoms and the probability of medical release (Odd Ratio: OR). The level of statistical significance was reached when p < 0·05·A value of *p* < 0·05 was considered significant. Calculations were performed using STATA 11.2.

## 3. Results

### 3.1 Demographic profile

In total, n = 172 were recruited for this study. The subjects were divided into experimental (EG: n = 110; median = 40·0 years old, min = 18·0, max = 75·0, 56F) and control groups (CG: n = 62; median = 37·5 years old, min = 18·0, max = 71·0, 34F). Table 1 shows the demographic profile and descriptions of comorbidity for the experimental and control group.

**Table 1.** Demographic Profile

| Variables | Experimental Group<br>(n= 110) | Control Group<br>(n= 62) | *p* |
| --- | --- | --- | --- |
| **Demographic profile** |  |  |  |
| Median Age (in years) | 40 | 37.5 |  |
| Interquartile Range (IQR) | [IQR<sub>25</sub>: 19; IQR<sub>75</sub>: 53] | [IQR<sub>25</sub>: 31; IQR<sub>75</sub>: 49] |  |
| **Gender - n°. (%)** |  |  |  |
| Female | 56 (50.91%) | 34 (54.84%) |  |
| Male | 54 (49.09%) | 28 (45.16%) | 0.62 |
| **Co-morbidities - n°. (%)** |  |  |  |
| HTA | 14 (12.73%) | 5 (8.06%) | 0.35 |
| DBT | 9 (8.18%) | 2 (3.23%) | 0.20 |
| Obesity | 8 (7.27%) | 1 (1.61%) | 0.11 |
| >60 years | 12 (10.91%) | 5 (8.06%) | 0.55 |
| Asthma | 1 (0.91%) | 0 (0.00%) | 0.45 |

HTA: Hypertension; DBT: Diabetes; Asthma. (\*) $p < 0.05$ .

Note, Table 1, that there is no statistical difference in demographics, gender, and comorbidities. Both samples are homogeneous (p > 0·05). Only, it was observed that the HTA, DBT, and obese population is greater in the EG than in the CG, a relationship 14:5, 9:2, and 8:1, respectively, with *p* > 0·05. Similarly, all were diagnosed with positive RT-PCR.

### 3.2 Decreased number of symptoms at 5th - 9th day, and posterior

Figure 2 shows as the first outcome the percentage of participants with symptoms reported from 5^th^ to 9^th^ day, which presented a greatest decrease in EG (48/98) when compared to CG (22/27) (*p* = 0·0026). After the date reported, from 10^th^ to 14^th^ day the medical examination did not show a significant difference in both groups.

**Figure 2.**
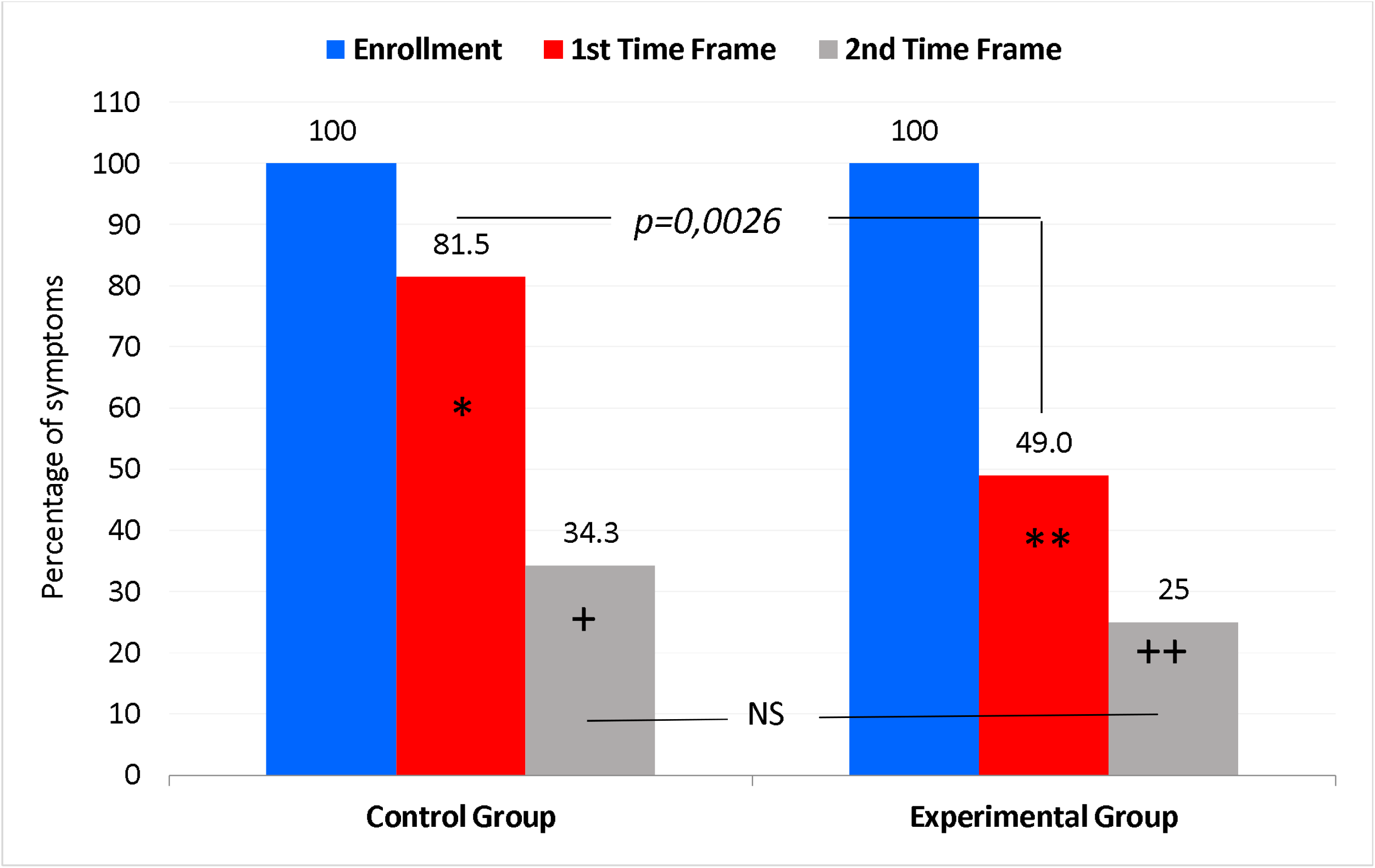
Percentage of number of symptoms: i) Enrollment, ii) 1^st^ time frame from 5^th^ to 9^th^ day: (*) *p* = 0·000 in CG, and (**) *p* = 0·000 in EG. iii) 2^nd^ time frame from 10^th^ to 14^th^ day: (+) *p* = 0·0007 in CG, and (++) *p* = 0·0187 in EG. NS: Non significant difference.

The association test between the sex variable in both groups showed that there are no significant differences in 1^st^ time frame and 2^nd^ time frame. The same happened when the age variable was analyzed.

Table 2 shows the clinical profile of COVID-19 symptoms in the 1^st^ time frame. The symptoms were divided in two categories: I) systemic symptoms, and II) upper airways symptoms.

**Table 2.** Percentage of number symptoms description 1^st^ time frame.

| Symptoms | Control Group |  |  | Experimental Group |  |  |
| --- | --- | --- | --- | --- | --- | --- |
|  | Enrollment | 1st Time Frame | *p* | Enrollment | 1st Time Frame | *p* |
| *I. Percentage Systemic Symptoms in COVID-19* |  |  |  |  |  |  |
| Fever | 46·8 | 1·6 | 0·0000 | 55·5 | 4·5 | 0·0000 |
| Diarrhea | 21·0 | 6·5 | 0·02 | 21·8 | 9·1 | 0·009 |
| Polymyarthralgia | 9·7 | 0·0 | * | 40·9 | 7·3 | 0·0000 |
| Headache | 32·3 | 11·3 | 0·005 | 43·6 | 7·3 | 0·0000 |
| Body pain | 32·3 | 9·7 | 0·002 | 17·3 | 2·7 | 0·0003 |
| Abdominal pain | 6·5 | 0·0 | * | 4·5 | 4·5 | 1 |
| Dyspnea | 9·7 | 3·2 | 0·14 | 6·4 | 5·5 | 0·77 |
| Tiredness | 12·9 | 1·6 | 0·01 | 8·2 | 3·6 | 0·15 |
| *II. Percentage Upper airway symptoms in COVID-19* |  |  |  |  |  |  |
| Taste and/or smell disturbance | 40·3 | 24·2 | 0·05 | 34·5 | 9·1 | 0·0000 |
| Odynophagia | 27·4 | 3·2 | 0·0002 | 15·5 | 0 | * |
| Cough | 21·0 | 17·7 | 0·6 | 19·1 | 10 | 0·05 |

Note in Table 2 the decrease in the percentage of systemic and upper airways symptoms reported in the medical examination in both groups. It was observed in the systemic symptoms that in both groups there is a favorable response to conventional treatment and to ivermectin treatment. However, ivermectin treatment was found to be more effective in alleviating upper airway symptoms, with a significant drop in cough (*p* < 0·05). With regard to odynophagia, and taste and / or smell disorder, the proportion test showed that the difference in symptom reduction was in favor of EG (*p* = 0·0004 and *p* = 0·00001, respectively).

Considering the 8-points ordinal scale used by WHO for categorize clinical improvement for COVID-19, mild patients are assigned to score 1 and 2, which means symptomatic patients no limitation of activities, and symptomatic patients with limitation of activities, respectively. Symptoms considered limiting because they need medical assistance are chest pain, conjunctivitis, lightheadedness, lumbago, respiratory symptoms, nausea and vomiting and imaging of lung involvement. Clinical examination was performed at the time of enrollment and was repeated between days 5^th^ and 9^th^ in both groups to determine what percentage of patients remained with these symptoms. We found in the 5^th^ to 9^th^ day that 49·9% of CG patients remained in score 2 from the scale, versus a 30% in EG, with a significative difference between both groups *p* = 0·039. Outpatients treated with ivermectin managed to climb a point on the scale, which translates into return to normal activity between 5^th^ and 9^th^ days.

### 3.3 Medical Release at 28 days after enrollment

The second finding of this study was the number of patients who received medical release at 28 days after enrollment (Figure 3) reaching category 0 in the scale.

**Figure 3.**
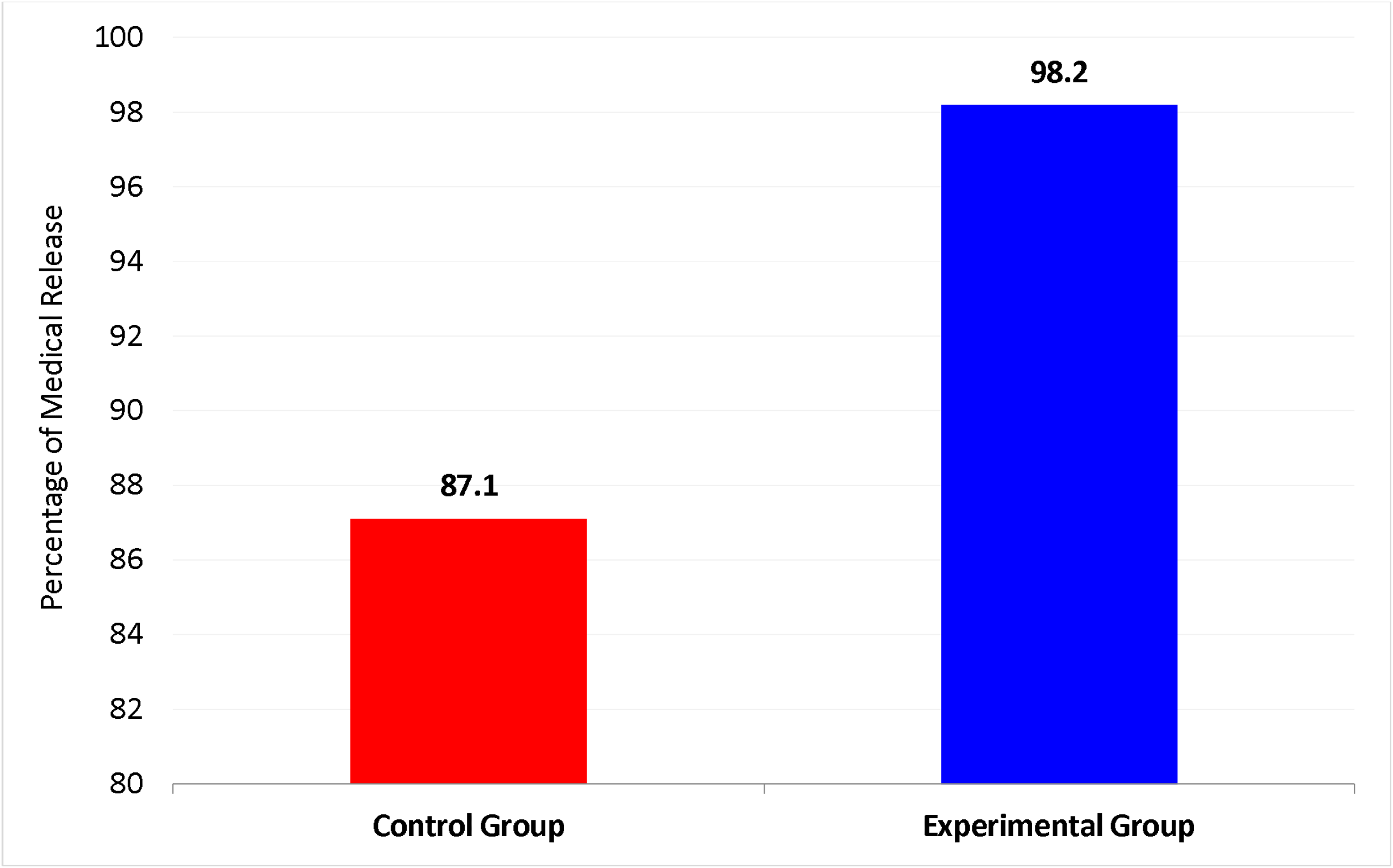
Medical Release at 28 days after enrollment

Because of the treatment, the EG had a higher medical discharge than the CG. Proportion was EG: n = 108/110; CG: n = 54/62. (*) *p* = 0·003. It was observed that there are no differences in achieving medical discharge when considering the age of the patient, both for the median age and for the distribution of interquartile. Similar results were observed for sex distribution.

### 3.4 Bivariate analysis and Logistic regression

Bivariate analysis showed 8 times more chance of receiving medical release in EG than CG (OR = 7·99, 95%, 1·64-38·97, *p* = 0·003). The effect of treatment with ivermectin to obtain medical release was analyzed by the logistic regression model based in the following variables: sex, age, and comorbidities. Then, the chance to obtain medical release was maintained in EG (OR= 10·37, CI = [2·05, 52·04]; *p* = 0·005) (Table 3).

**Table 3.**
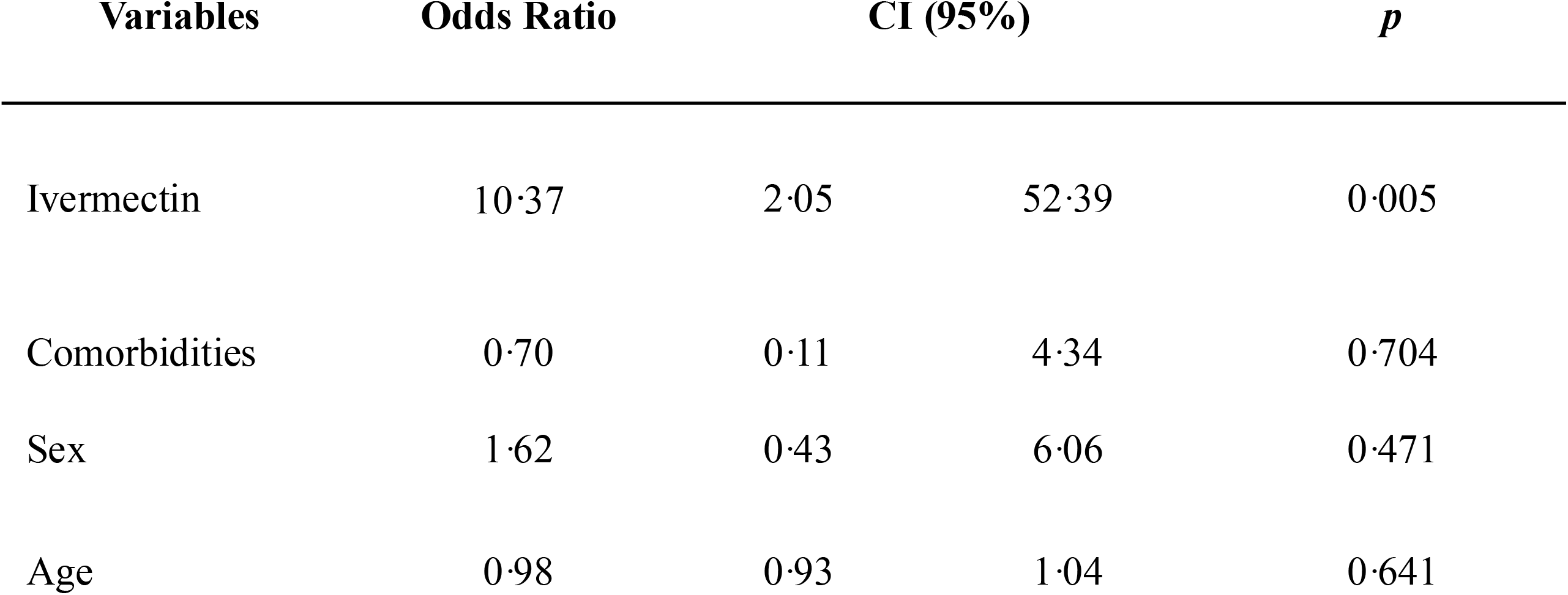
Logistic regression

## 4. Discussion

This study was designed to evaluate the potential of ivermectin as a repositionable drug, for treatment of mild cases of COVID-19. According to outcomes, ivermectin has shown effect in clinical manifestations of COVID-19, like decrease in the percentage of symptoms reported in the medical examination in both groups, and an increase of chances for medical release. Additionally, the treatment with ivermectin is early in relation to the onset of symptoms and diagnosis by RT-PCR (2 days in both groups). Currently, many studies about ivermectin and its potential against SARS-CoV-2 are complete or in development. A pilot study about effects of ivermectin in treatment for early COVID-19^17^, sheds some light on the potential mechanism of action of ivermectin against the virus, but in the trial, ivermectin has not shortened the duration of symptoms as fever or malaise, which are associated with systemic inflammation. That is contrast with the reports of IVER-Leve study, in which outpatients report a significant drop in the percentage upper airway symptoms in COVID-19 (taste and/or smell disturbance, odynophagia, cough), see Table 2.

Concomitantly, the results reported here show that the use of ivermectin produces a decrease in number of symptoms reported by patients, such us feverish and diarrhea, but above all, a significant decrease in taste and smell lost, which is related to the effects of viral load on upper air vials in patients with mild COVID-19. Other percentages of patients with symptoms that decreased were polyarthralgia, headache and abdominal pain (p < 0·05). Chaccour et al., reported in the patients treated with ivermectin a significant diminution of 50% less anosmia/ hyposmia than those in the placebo group (76 vs 158 patient-days of anosmia / hyposmia)^17^. The ivermectin group also reported 30% fewer coughs (68 vs 97 patient-days of cough). However, in this study there were no major differences between ivermectin and placebo in the reported patient days of fever, general malaise, headache or nasal congestion. No patient from either group progressed to severe disease^17^.

Recently, Lopez Medina et al. published a study of ivermectin in relation to the time resolution of symptoms in mild patients^18^. Although they do not recommend the use of ivermectin as a treatment for COVID-19, there are some coincidences (and differences) with the results of the present study. We agree that there are not significant differences between both groups at 14 and 21 days (see Figure 2), but our main finding shows that the effect of treatment is observed between 5 and 8 days after the patient starts treatment. This difference may be due to the administration of the dose, which in our case is weekly and not daily, as in the work by Lopez Medina et al.^18^

Another difference pointed out in our study is related to the clinical follow-up of the patients, which in our study was carried out in person in the Primary Health Centers, while in Lopez Medina it was carried out by telemedicine. In this sense, virtual monitoring is less accurate in relation to the recording of symptoms.

Even if the sample of the studies is small to make solid conclusions, there are results which provide evidence of the potential benefit of an early intervention with ivermectin for the treatment of patients diagnosed with mild stages of COVID-19^19^. There are many studies that present a viral load reduction, as has been suggested by Caly et al. in vitro. This could have the potential effect on disease progression and spread ^9,17,19,20^.

A study performed in 167 patients with mild to severe COVID-19 from Argentina, found that none of the mild or moderate cases of COVID-19 who received the experimental treatment with ivermectin were hospitalized, and only one patient died (0.59%)^21^. In México a comparative effectiveness study was performed among patients with laboratory confirmed SARS-CoV-2 infection. The experimental group receive a TNR4. TNR4 consists of four drugs administered orally to COVID-19 cases with mild or moderate symptoms: (1) ivermectin, 12 MG single dose; (2) Azithromycin 500 mg for 4 days; (3) Montelukast, 60 mg on the first Day and then 10 mg between days 2 to 21; and (4) acetylsalicylic acid, 100 mg for 30 days. This study indicated that the TNR4 significantly increases the likelihood of a full recovery within 14 days after the onset of symptoms, and decreases the risk of hospitalization or death among ambulatory cases of COVID-19^22^.

Addition of ivermectin to standard care is an effective drug for treatment of COVID-19 patients with significant reduction in mortality and hospital stay days compared to Hydroxychloroquine plus standard treatment only. Early use of ivermectin is very useful for controlling COVID-19 infections, improving cytokines storm and prophylaxis of frontline health care as well as household contacts^23^.

Our study has sizable limitations. The absence of a placebo group is due to the lack of funding from a sponsor and the need to guarantee treatment to the entire population. Other way, sample size, although representative, is small to obtain conclusive results. It’s a descriptive study of clinical follow up at 28 days. However, shows overlaps in benefits with other authors, and taking together, these results are encouraging for further study about repurposing ivermectin for the treatment of COVID-19, considering that is an inexpensive drug and is accessible in the local pharmaceutical industry (Argentina).

We suggest new clinical intervention studies in our region and partners in other countries that may show the effect of the IVER compound in mild-stage outpatients.

## 5. Conclusion

Treatment with ivermectin in the population of outpatients with mild stage COVID-19 disease managed to significantly reduce the number of symptoms on days 5 and 9. Subsequent medical examination did not show significant differences.

This treatment also had a significantly effect (p=0·003) in achieving medical release at EG vs in CG.

The treatment with ivermectin could significantly prevent the evolution to serious stages since the EG did not present any patient with referral to critical hospitalization. In both groups, the patients did not advance to highest scores in the ordinal scale, which represents more compromised stages of the disease.

This proposed treatment brings additional benefits in relation to the improvement in the patient’s clinical condition, without this having an impact on adherence. It is also added that it did not produce lack of adherence or adverse effects.

We found that patients who took ivermectin had a greater than 89·1% probability of being released at the end of the intervention than the control group. This probability remains even in association with the presence of comorbidities.

No deaths were recorded in any group of these mild patients. It should be noted that the median age (40-year-old) and the presence of comorbidities were similar in both groups.

## Authors’ contributions

ESO supervised the database. GGB, ESO and DGG contributed with the data processing and contributed to the statistical analysis. ESO, DGG and MPB were responsible for writing the manuscript. TM, YB and PT contributed to data collection. REC and LMR were the institutional managers to carry out the work. MPB supervised the project.

## Data Availability

Data Sharing Statement
Data available:Yes
Data types:Participant data with identifiers
How to access data:E-mail request for data: - Dr. Maria Peral de Bruno
Who can access the data:Researchers whose proposed use of the data has been approved
Types of analyses:for a specified purpose
Mechanisms of data availability:with investigator support

## Acknowledgements

All the authors are grateful for the collaboration of the health and administrative personnel of the Primary Cares Centers: Adolfo de la Vega, San Martín, 25 de Mayo, Alberdi, Alderete, Nuestro Sr. del Milagro, Vial III, Delia Fernandez Palma, Esquina, Fernando P. Riera, Bella Vista Hospital, Los Ralos Hospital, Lomas de Tafí, Los Pocitos, Villalonga Policlinic, Santa Ana, and Active Search Center for Febrile and Symptomatic (in Spanish language: *”Centro de Búsqueda Activa de Febríles y Asintomáticos”)*. The authors thank Ms. Emma Lis Garat (Dirección Investigación en Salud, SI.PRO.SA, Tucumán, ARG) and Dr. Augusto Bellomio (INSIBIO UNT, Tucumán, ARG) for critical reading of the manuscript.

